# Effect of children’s BMI on the association between parents’ socio-demographic and economic characteristics, and Parental perceptions of their child’s weight status in South-western Sydney: The Growing Healthy Kids Survey, Australia

**DOI:** 10.1101/2024.07.02.24309846

**Authors:** Vilas Kovai, Shanley Chong, Bin Jalaludin, Janice Tang, Michelle Camilleri, Margaret Thomas, Francis Fox, Mandy Williams

## Abstract

**Background:** Parents’ perception of children’s weight status has been recognised as one of the influencing factors when planning childhood overweight and obesity prevention programs. We aimed to examine whether children’s BMI modified the association between parents’ socio-demographic and economic characteristics and their perceptions of their children’s weight status in South Western Sydney Local Health District.

**Methods:** We analysed 1,353 parents’ perceptions of their child’s weight status using the ‘Growing Healthy Kids-Population Health Survey’ baseline data. Generalised linear multinomial mixed models were used to determine the associations. Results are presented as odds ratio and associated 95% confidence intervals by BMI.

**Results:** This study found that the weight status of about 23% of healthy-weight children, 65% of overweight children and 55% of obese children was incorrectly perceived by parents. Father as the primary parent and parents who completed ≤12 years of school education were less likely to perceive their overweight or obese children as overweight. Parents who lived in more disadvantaged areas were more likely to perceive their obese children as overweight. Parents who lived in moderate disadvantaged areas were less likely to perceive their overweight children as overweight. Parents who had reported an annual household income between $52,000-$103,999 were more likely to perceive their overweight children as of healthy weight and parents who reported an annual household income below $103,999 were twice as likely to perceive their obese children as having a healthy weight.

**Conclusion:** The findings suggest that there may be benefit in designing interventions that focus on improving parents’ misperception of children’s weight status. The groups of parents who should be prioritised for preventive public health interventions to modify the parental misperceptions of their child’s weight status include a) fathers, b) parents who completed ≤12 years of school education and parents who had Trade, TAFE or Diploma as educational qualification, c) parents living in moderately disadvantaged areas and d) parents with low and high household-income.

## Introduction

Childhood overweight and obesity is a global public health priority ^1^. Childhood obesity is associated with an array of short and long-term effects including deteriorating health and well-being, bullying by peers, and emotional and mental health issues^2^. Australia has one of the highest rates of obesity in the world. In 2018, two-thirds of Australian adults and a quarter of Australian children aged 5-17 years were overweight or obese^3^. Of particular concern is that Australians live on average 2.7 years less due to overweight and obesity^1^. Additionally, children who are overweight or obese are more likely to remain overweight or obese in adulthood^4^. Hence, the prevention of childhood obesity is of utmost importance.

Parents’ misperception (defined as the discrepancy between measured weight status and subjective assessment of children’s weight status by parents) about their children’s weight status has been recognised as one of the influencing factors when planning childhood overweight and obesity prevention programs^5^. However, less than 50% of parents accurately perceive their child’s overweight or obese status^6–8^ and recent New South Wales (NSW) data shows that three in four parents of overweight children and nearly one-third of parents of obese children believed their child was about the right weight^9^. While there has been research investigating the association between demographic factors including socioeconomic status (SES) with parental misperception of their child’s weight status, the findings are inconsistent. Evidence from a meta-analyses^8^ shows that children’s age and weight or body mass index (BMI) are important moderators of the parent’s underestimation of their child’s overweight status and gender, parent’s weight and method of perception (visual versus non-visual) were important moderators of parents underestimation of child’s weight status.

A meta-synthesis of parental perceptions about a child’s weight status^10^ reported that parents are more likely to misperceive a child’s weight if they are overweight than if the child is of normal weight, regardless of the child’s age. The meta-synthesis also showed that more mothers compared to fathers, and mothers with lower education, lower household income, and who hold size-related cultural beliefs (e.g., acceptance of slim daughters and larger body size for boys) are factors associated with parents’ underestimation of child’s overweight or obese status. Parents were more likely to overestimate girls’ weight compared to boys^10^.

In a recent systematic review of studies investigating parents’ perception of their children’s weight, nearly half of the studies found no association between SES and parental misclassification of children’s weight, while others showed mixed results^11^. Parents’ correct identification of their children as overweight and obese is an important aspect of childhood obesity prevention and management^12, 13^. Parents who acknowledge and are concerned about their children’s weight will be more likely to implement effective changes to their children’s lives and regulate weight gain/reduction^14^.

Misperception of children’s weight is associated with a lower likelihood that parents intend to take action to help their children lose weight^15^. Parental misperception may lead to unhealthier diets and food choices for their children^16–18^. Children whose parents misperceive them to be overweight have a negative view of their body size and actively try to lose weight compared to their peers^19^. However, parental misperceptions of being overweight can be stigmatising and actively harmful in the long term because body dissatisfaction and associated dieting intentions among adolescents are predictors of future weight gain^20, 21^.

Although multiple factors are associated with parental underestimation of overweight and obesity status, to our knowledge, there are no population-level studies in Australia that comprehensively examined the child’s BMI as an effect modifier on the relationship between parents’ characteristics and the accuracy of parental perception of their child’s weight status. Given the high prevalence of childhood overweight and obesity in the state of NSW, Australia^22^, and our findings that 37% of children in the South Western Sydney Local Health

District (SWSLHD) are overweight or obese^23^, a greater understanding of population-level parental socio-demographic factors including socioeconomic gradient in parental perceptions of child’s weight would be useful to guide state-wide and local health promotion programs that aim to achieve healthy weight among children.

SWSLHD has an estimated population of over 1 million living in its seven Local Government Areas which include more densely populated areas of Bankstown, Fairfield, Liverpool and Campbelltown and less densely populated areas of Camden, Wollondilly and Wingecarribee^24^. An estimated 21% of residents are under 15 years of age and a further 15% are in the 15-24 year age range^24^. SWSLHD also has the largest population of culturally and linguistically diverse (CALD) communities in NSW, with 42% of residents born overseas, about 51% of families speaking only English at home and approximately 50% of persons aged over 15 years earning < AU$650 per week^24^. Overall, SWSLHD provides health services to a rapidly growing culturally and linguistically diverse population across a geographical area of approximately 6,243 square kilometres^25^. These demographic features and high rates of childhood overweight and obesity make SWSLHD an ideal location for this study. This study examined whether children’s BMI modified the association between parent’s characteristics (parent’s role as mother or father, parent’s country of birth (COB), parent’s level of education and parent’s SES) and parental perceptions of their children’s BMI (overweight, obese, and healthy weight).

## Methodology

### Ethics Approval & Formal Consent

This study was approved by the Human Research Ethics Committee of South Western Sydney Local Health District (HE18/078). Informed verbal consent was obtained from all the study participants (i.e., Parents or carers aged ≥18 years who could provide the maximum information about the selected child) before the interviews. The data was collected from October and November 2018. The data was accessed for research purposes from February 2021 and the authors had no access to information that could identify participants during or after data collection.

### Sample

We analysed baseline data from the ‘Growing Healthy Kids-Population Health survey (GHK-SWS survey, 2018) for this study. The GHK-SWS baseline study—a cross-sectional population-based survey of parents or carers of children aged 5-16 years in southwestern Sydney, NSW, Australia.

The main paper from the GHK-SWS study presents a detailed description of the study design and methodology^26^. In brief, the GHK-SWS baseline survey was developed using validated questions from the 2018 NSW Population Health Survey^27^, NSW Schools Physical Activity and Nutrition survey^28^ and the Make Healthy Normal campaign evaluation surveys^29^. The baseline survey also included new questions when no other suitable questions were available. The survey domains included: socio-demographics, health status, parental perception of their child’s height and weight, consumption of fruit, vegetables and discretionary foods, physical activity participation and related behaviours, and several knowledge and attitudes questions. The questionnaire was translated into Arabic, Vietnamese, and Cantonese/Mandarin (simplified Chinese) as these are the major language groups in SWSLHD. Accredited translators and the GHK team conducted translations and the back translations to ensure the use of culturally appropriate language by the interviewers. GHK-SWS study data was collected using a computer-assisted telephone interview (CATI) system.

The target population included all households within SWSLHD with children aged 5-16 years. The sample frame consisted of both landline and mobile telephone numbers of residents. An age and sex-matched sample size of 1,815 respondents was recruited and included in the baseline survey. This was based on a power of 0.8 with an alpha of 5% and 28.4% baseline estimated prevalence^23^ of childhood overweight and obesity in residents of south western Sydney.

The primary caregiver (parent/carer who could provide maximum information about the selected child) was interviewed from each randomly selected household. Interviewers collected information on the number of eligible children living in the household and their sex and ages. If the household had more than one eligible child, the interviewer randomly selected one of the eligible children using the CATI system’s random number generation. If the respondent of the selected household was not available after five calls, the interviewers recorded this as ‘household member not available’.

## Measures

### Outcome measure

This study’s main outcome measure was ‘parental perceptions of their child’s weight status’ as used in the NSW Schools Physical Activity and Nutrition Study^9^. Parents were asked to select a response to the question “Do you consider your child to be: very underweight, slightly underweight, at a healthy weight, slightly overweight, very overweight, or not sure”. When parents reported their child as slightly overweight, we classified this as a perception of the child being overweight, and if the child was reported as very overweight, we classified this as a perception of the child being obese. Children reported as underweight (both self-reported and perceived) were excluded from the study as the focus of the study was to understand parental perceptions that their slightly overweight or very overweight child was of a healthy weight.

### Predictors of interest

The study’s main predictors of interest were parent’s characteristics and SES at both the household level and area-level. In this paper, parent refers to either mother (73% of respondents) or father (27% of respondents) and only their responses to survey questions were included in the analysis. Hence, data from other carer relationships (e.g., grandparents or relatives) were excluded from the analysis (n=51), which also makes our data comparable with the other studies that used only parents’ data. Parent’s characteristics include the parent’s relationship with the child (mother or father), parent’s country of birth (Australia-born and overseas-born), parent’s educational attainment 1: ≤Year 12 of education, 2: Trade, Technical and Further education (TAFE), or Diploma, and 3: University degree).

SES was assessed by two measures: annual household income (before tax), and area-level SES. Annual household income was grouped into four categories: $0-$51,999, $52,000-$77,999, $78,000-$103,999 and $104,000 or more.

Area-level SES was measured using the 2016 Index of Relative Socio-Economic Disadvantage (IRSED) at the postcode level ^30^. The IRSED scores were created by the Australian Bureau of Statistics to compare social and economic disadvantage across geographic areas in Australia. The index is derived from census variables such as income, educational attainment, unemployment, and people working in unskilled occupations. IRSED at postcode level was categorised into tertiles (tertile 1: most disadvantaged/low SES, tertile 2: moderately disadvantaged/moderate SES, and tertile 3: least disadvantaged/high SES).

### Effect modifier

We tested whether children’s BMI status modified the associations between parent’s socio-demographic characteristics and the perception of their child’s weight status. Children’s BMI was categorised using the age and sex-specific International Obesity Task Force (IOTF) definitions ^31^: thin (BMI<18 kg/m^2^), healthy weight (18 kg/m^2^ ≤BMI<25 kg/m^2^), overweight (25≤BMI<30 kg/m^2^) and obese (BMI≥30 kg/m^2^). To determine BMI, participants were asked two questions: 1. How tall is your child without shoes? (in centimetres). 2. How much does your child weigh without clothes or shoes? (in kilograms).

### Mediators

The following three parents’ perceptions about childhood obesity and their child’s diet were explored as mediators (see figure 1): a) It’s alright for a child aged 5-16 years to be overweight; b) Drinking sugar-sweetened soft drinks too often as the cause of overweight and obesity in children; and c) Overall, do you think your child’s diet is healthy? Parent’s agreement with each of the first two statements was measured on a 5-point Likert scale (from strongly agree to strongly disagree) and grouped into three categories: agree (strongly agree and somewhat agree combined), neither, and disagree (strongly disagree and somewhat disagree combined). The third statement was measured on a 4-point Likert scale and was grouped into two categories: yes (‘Yes, always’ and ’Yes, mostly’ combined) and no (’No, not really’ and ‘Not at all’ combined).

**Figure 1:**
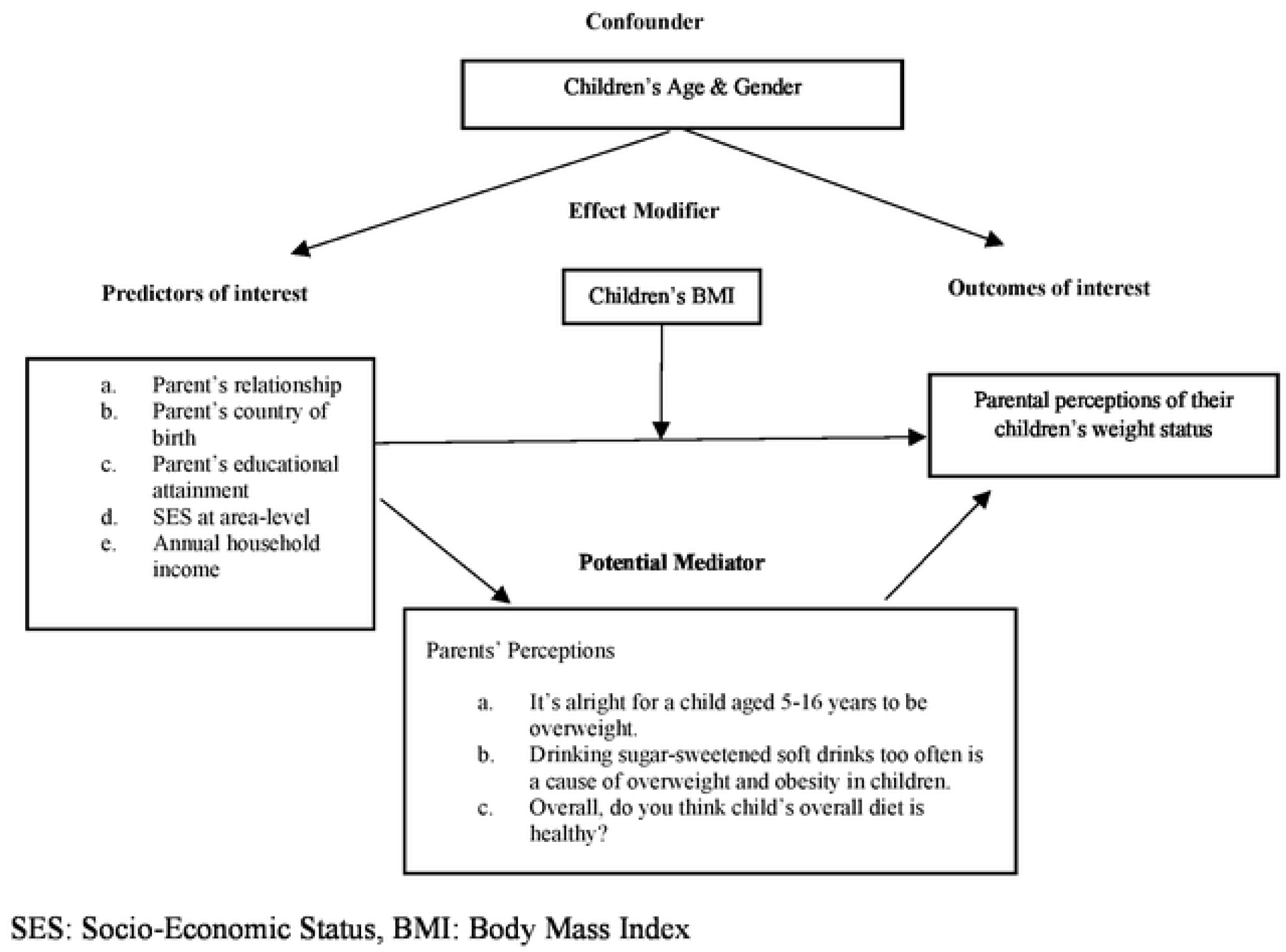
A directed acyclic graph (DAG) showing potential confounders, mediators, and effect modifiers in the relationship between predictors of interest and parental perceptions of their children’s weight status.

### Data analysis

A directed acyclic graph (DAG) was used to identify potential confounders, mediators, and effect modifiers in the relationship between parent’s characteristics, SES, and parental perceptions of their child’s weight status (Figure 1).

Following a comprehensive descriptive analysis, the R package “mma” was used to conduct multiple mediation analysis^32^. Multiple mediation analysis examined the presence of three potential mediators: parents’ perception of overweight and obesity, children’s overall diet, and child’s eating behaviour. The algorithms for the estimation were repeated 50 times with 500 times of bootstrap resampling. Perception of overweight and obesity, children’s overall diet, and children’s eating behaviour were not significant mediators and were included as confounders in our final model. Proc Glimmix procedure in SAS v9.4^32^ was then used to conduct generalised linear multinomial mixed models together with random intercept accounting for Local Government Area clustering effects to determine associations between area disadvantage and parental perceptions of their child’s weight status. Parental perceptions of their child’s weight as healthy were used as the reference group. Children’s age and gender were included as confounders in the final model.

Effect modification by child’s BMI was tested and were all statistically significant (p<0.0001). Results from the multinomial multilevel regression model are presented as odds ratios (ORs) and associated 95% confidence intervals (95% CIs) by BMI category. Survey weighting was used in all analyses to ensure the survey sample was representative of all children living in SWSLHD.

## Results

The sociodemographic and other characteristics of participants are presented in Table 1. After the exclusion of other caregivers (n=51), underweight children (n=224), and children perceived as very underweight and slightly underweight (n=254), the study findings are presented from the analysis of 1,353 children. Of the total study sample, 56% were aged 5-11 years with a mean of 11.0 years (SD=0.12), and 50% were girls. Seventy-three per cent of parents were mothers. About half of the parents were born in Australia (51%), approximately one-third had completed a university degree, and another one-third had completed Technical and Further Education (TAFE) or a diploma degree (33%). About one-third had reported an annual household income of less than AUD 52,000. Fifty-five percent of participants lived in the most disadvantaged areas and 26% lived in the least disadvantaged areas.

**Table 1:** Descriptive statistics of child’s and parent’s characteristics, SES at household level and area-level, health knowledge and outcome variable, n=1353

| Variable | Level | n | % |
| --- | --- | --- | --- |
| Age | 5-11 years | 761 | 56.1 |
|  | 12-16 years | 592 | 43.9 |
| Gender | Boys | 683 | 50.1 |
|  | Girls | 670 | 49.9 |
| Parent | Mother | 964 | 73.2 |
|  | Father | 389 | 26.8 |
| Parent's country of birth (missing=45) | Australia-born | 695 | 51.0 |
|  | Overseas born | 658 | 49.0 |
| Parent's educational attainment<br>(missing=6) | <=Year 12 | 372 | 32.2 |
|  | Trade, TAFE, or Diploma | 467 | 33.0 |
|  | University degree | 508 | 34.9 |
| Annual Household Income-before tax<br>(missing=203) | \$0-\$51,999 | 306 | 33.1 |
| | \$52,000-\$77,999 | 223 | 21.2 |
| | \$78,000-\$103,999 | 222 | 16.5 |
| | \$104,000 or more | 399 | 29.3 |
| Area – Level SES | Most disadvantaged | 740 | 54.5 |
|  | Middle disadvantaged | 242 | 17.5 |
|  | Least disadvantaged | 371 | 26.0 |
| BMI | Healthy weight | 743 | 55.1 |
|  | Overweight | 347 | 24.4 |
|  | Obese | 263 | 20.5 |
| Health knowledge |  |  |  |
| Child's overweight is all right (missing=2) | Agree | 178 | 13.4 |
|  | Neither agree nor disagree | 95 | 7.5 |
|  | Disagree | 1078 | 79.1 |
| Drinking sugary drinks too often is a cause of overweight/obesity among children (missing=4) | Agree | 1206 | 89.9 |
|  | Neither agree nor disagree | 47 | 3.0 |
|  | Disagree | 96 | 7.2 |
| My child's overall diet is healthy (missing=15) | No | 165 | 11.5 |
|  | Yes | 1173 | 88.5 |
| Outcome variable |  |  |  |
| Parent's perception of child's weight<br>(missing=2) | Healthy | 1038 | 77.1 |
|  | Slight and very overweight | 315 | 22.9 |

SES: Socioeconomic status
BMI: Body Mass Index
TAFE: Technical and Further Education

On parent-reported height and weight, more than half of the children (55%) were within the healthy weight range, 24% were overweight, and 21% were obese. Most parents perceived their children as having a healthy weight (77%), and 23% perceived their children as overweight or obese. Most parents disagreed that being overweight is all right (79%), agreed that drinking sugary drinks too often is the cause of being overweight/obese (90%), and reported their child’s overall diet to be healthy (89%) (Table 1).

Overall, 65% of parents (914/1353) perceived their children’s weight correctly and 35% perceived their children’s weight incorrectly. The weight status of about 23% of healthy-weight children, 65% of overweight children and 55% of obese children was incorrectly perceived by parents. Overall, the sensitivity of the overweight category was 79.4% (95% CI=79.1%-79.7%), and the specificity was 37.1% (95% CI=36.5%-37.6%). The sensitivity of the obese category was 85.1% (95% CI=85.1-85.3%), and the specificity was 40.0% (95% CI=39.4%-40.6%). Finally, the sensitivity of the healthy weight category was 77.1% (95% CI=72.5%-81.8%), and the specificity was 64.6% (95% CI=61.7%-67.6%).

Table 2 (unadjusted model) presents the associations between child’s and parent’s characteristics, SES at both the household level and the area level, parents’ perceptions about childhood obesity and child’s diet and parents’ perception on their children’s weight from single variable models.

**Table 2:** Associations between child’s and parent’s characteristics, SES at household level and area-level, health knowledge and parental perceptions on their children’s weight by children’s BMI

|  |  | Parents perception of their child's weight status |  |  |  |  | Type 3, p-value |
| --- | --- | --- | --- | --- | --- | --- | --- |
|  |  | Healthy weight |  | Overweight or Obese |  | Unadjusted OR, 95% CI |  |
|  |  | n | % | n | % |  |  |
| Age | 5-11 years | 604 | 79.0 | 157 | 21.0 | 1 | <.0001 |
|  | 12-16 years | 434 | 74.6 | 158 | 25.4 | 1.35, 1.31-1.39 |  |
| Sex | Boys | 524 | 78.5 | 159 | 21.5 | 1 | <.0001 |
|  | Girls | 514 | 75.6 | 156 | 24.4 | 1.25, 1.21-1.28 |  |
| Parent relationship with child | Mother | 744 | 77.8 | 220 | 22.2 | 1 | 0.779 |
|  | Father | 294 | 74.9 | 95 | 25.1 | 1.05, 0.97-1.04 |  |
| Parent's country of birth | Australia-born | 549 | 79.7 | 146 | 20.3 | 1 | <.0001 |
|  | Overseas born | 489 | 74.3 | 169 | 25.7 | 1.15, 1.11-1.18 |  |
| Parent's educational attainment | <=Year 12 | 305 | 76.2 | 95 | 23.8 | 1.06, 1.02-1.10 | <.0001 |
|  | Trade, TAFE, or Diploma | 333 | 75.6 | 103 | 24.4 | 1.15, 1.11-1.19 |  |
|  | University degree | 392 | 79.1 | 115 | 20.9 | 1 |  |
| Annual household income | \$0-\$51,999 | 215 | 71.0 | 91 | 29.0 | 1.10, 1.06-1.14 | <.0001 |
| | \$52,000-\$77,999 | 181 | 83.4 | 42 | 16.6 | 0.59, 0.57-0.62 | |
| | \$78,000-\$103,999 | 175 | 78.8 | 47 | 21.2 | 0.81, 0.77-0.85 | |
| | \$104,000 or more | 307 | 76.2 | 92 | 23.8 | 1 | |
| SES | Most disadvantaged | 547 | 74.8 | 193 | 25.2 | 1.03, 0.98-1.08 | 0.0001 |
|  | Middle disadvantaged | 186 | 74.8 | 56 | 25.2 | 0.94, 0.89-0.99 |  |
|  | Least disadvantaged | 305 | 86.6 | 66 | 16.4 | 1 |  |
| BMI | Healthy weight | 671 | 90.5 | 72 | 9.5 | 1 | <.0001 |
|  | Overweight | 225 | 65.1 | 122 | 34.9 | 4.96, 4.77-5.16 |  |
|  | Obese | 142 | 55.2 | 121 | 44.8 | 6.94, 6.67-7.21 |  |
| Childhood overweight/obesity is a problem (missing=4) | Agree | 914 | 75.5 | 3015 | 24.5 | 1 | <.0001 |
|  | Neither agree nor disagree | 49 | 96.2 | 3 | 3.8 | 0.03, 0.02-0.42 |  |
|  | Disagree | 72 | 87.1 | 11 | 12.9 | 0.39, 0.36-0.43 |  |
| Child being overweight is all right | Agree | 112 | 62.3 | 66 | 37.7 | 2.30, 2.21-2.39 | <.0001 |
|  | Neither agree nor disagree | 67 | 73.5 | 28 | 26.5 | 1.48, 1.40-1.56 |  |
|  | Disagree | 857 | 79.8 | 221 | 20.2 | 1 |  |
| Drinking sugar sweetened soft drinks too often is a cause of overweight among children | Agree | 915 | 76.3 | 291 | 23.7 | 1 | <.0001 |
|  | Neither agree nor disagree | 42 | 82.0 | 5 | 18.0 | 0.75, 0.69-0.82 |  |
|  | Disagree | 77 | 83.0 | 19 | 17.0 | 0.74, 0.69-0.79 |  |
| My child's overall diet is healthy (missing=17) | No | 91 | 52.7 | 74 | 47.3 | 4.05, 3.89-4.22 | <.0001 |
|  | Yes | 938 | 80.4 | 235 | 19.6 | 1 |  |

SES: Socioeconomic status, BMI: Body Mass Index, TAFE: Technical and Further Education

Using parental perception of their child as a healthy weight (whether correct or not) as the reference group, parents with a female child and child aged 12-16 years were more likely to report them to be overweight or obese. Parents who were born overseas, parents who had not completed a university degree, parents who reported an annual household income under $51,999, and parents living in the most disadvantaged areas, were more likely to perceive their children as overweight or obese (Table 2).

Parents who agree childhood overweight and obesity is a problem, disagreed that being overweight in childhood is alright, agree that drinking sugary drinks too often is a cause of overweight/obesity among children, or thought their child’s overall diet was not healthy were more likely to perceive their child as overweight or obese (Table 2).

Figures 2 and 3 (adjusted models) present the associations between the main predictors of interest (i.e., children and parents’ characteristics and SES at household and area level) and perception of the child’s weight by parent-reported BMI weight status after adjusting for confounding.

**Figure 2:**
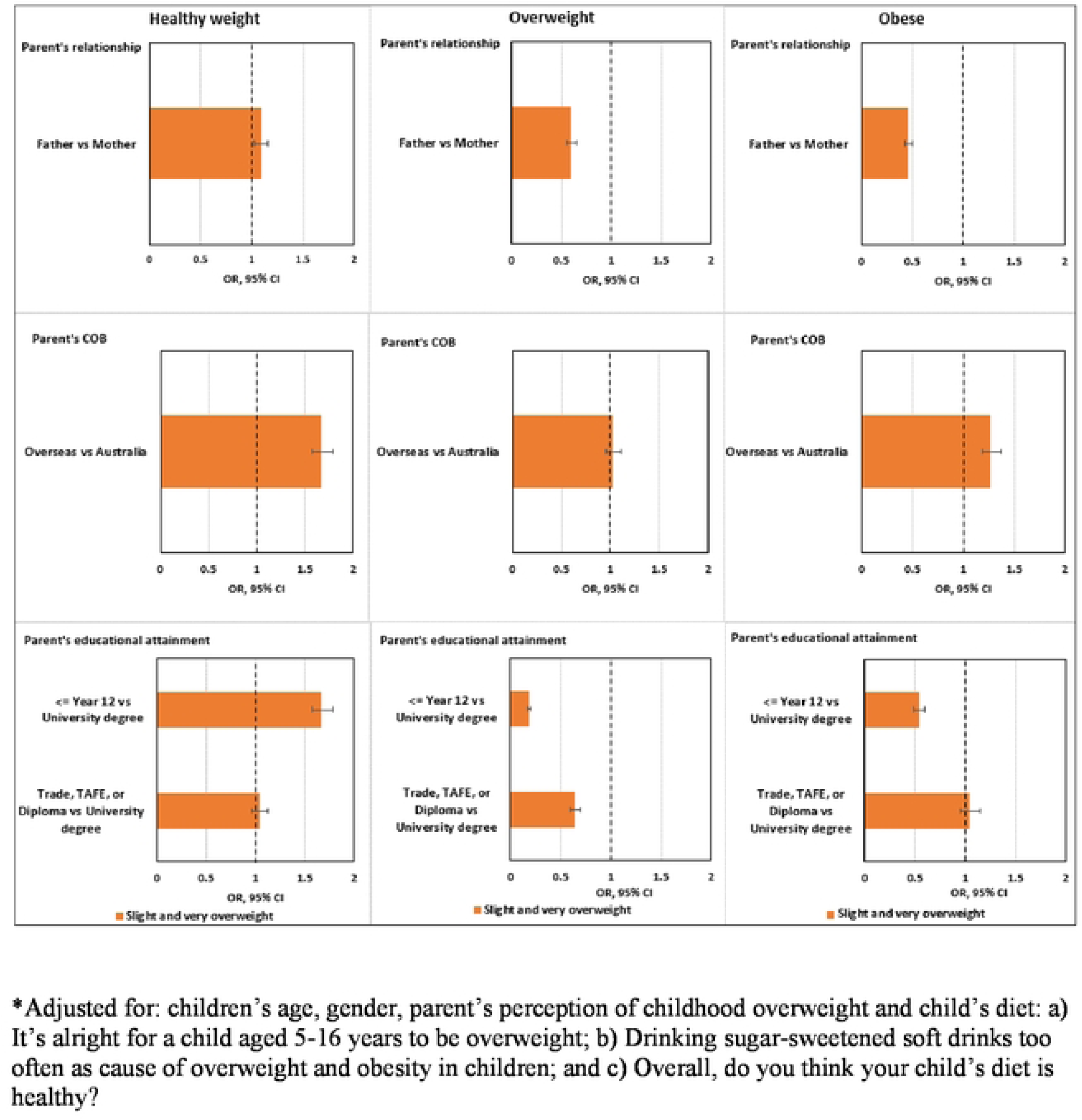
Associations between parent’s characteristics and perception of child’s weight by child’s BMI*.

**Figure 3:**
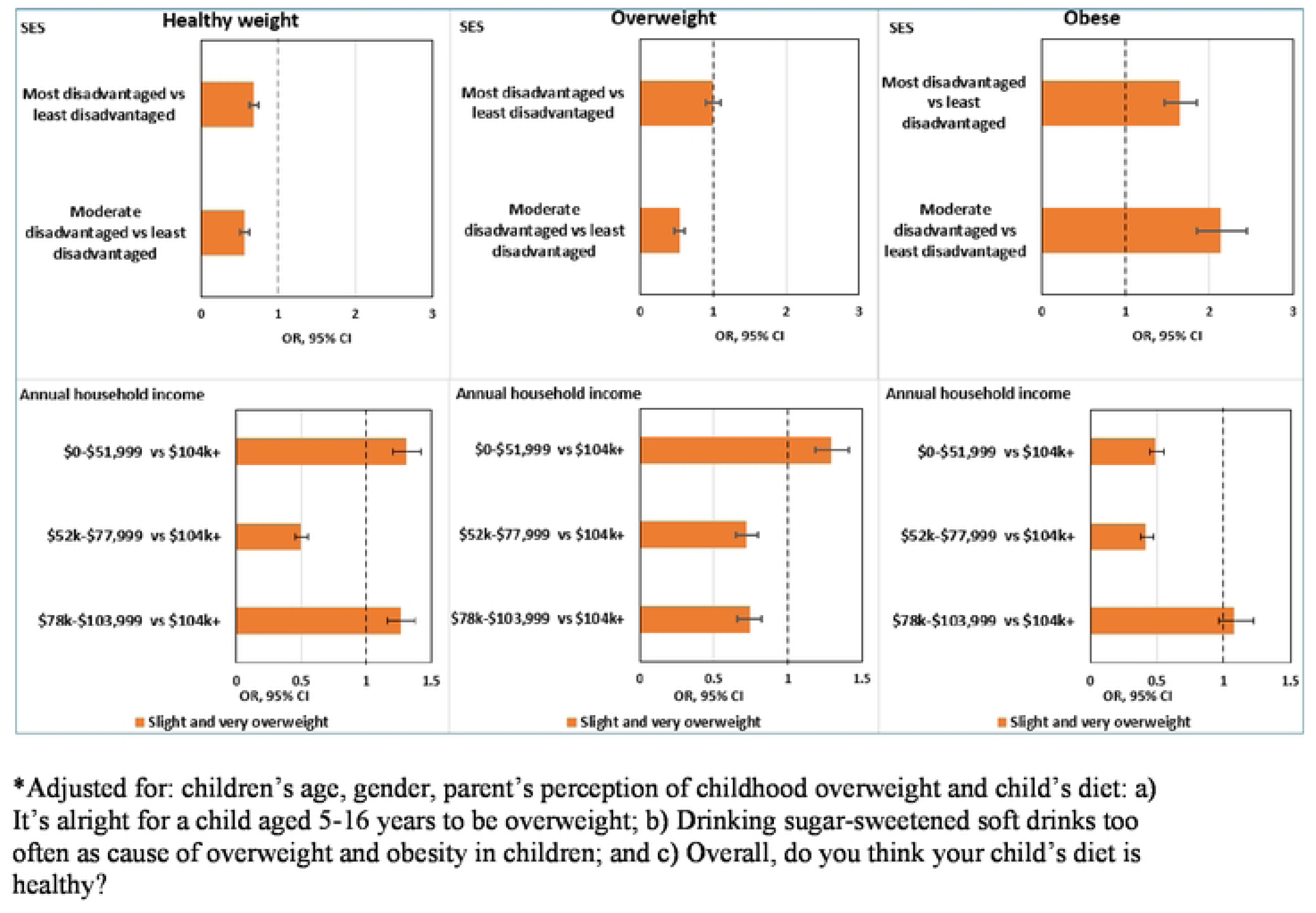
Associations between SES and Parent’s perception of child’s weight by child’s BMI.

Figure 2 shows that Father as the primary parent (overweight: OR=0.60, 95% CI=0.55-0.65; obese: OR=0.46, 95% CI=0.43-0.50) and parents who completed ≤12 years of school education (overweight children: OR=0.18, 95% CI=0.17-0.20; obese: children OR=0.54, 95% CI=0.49-0.459) were less likely to perceive their overweight or obese children as overweight or obese.

Overseas-born parents were more than twice as likely to perceive their healthy-weight children as healthy weight (OR=1.70, 95% CI (1.60 -1.80) and obese children as obese (OR=1.30, 95% CI (1.20 -1.40).

Figure 3 shows that parents who lived in more disadvantaged areas were more likely to perceive their obese children as overweight or obese (most disadvantaged areas: OR=1.65, 95% CI=1.46-1.86; moderate disadvantaged areas: OR=2.13, 95% CI=1.86-2.45). Parents who lived in moderate disadvantaged areas were less likely to perceive their overweight children as overweight or obese (OR=0.54, 95% CI=0.47-0.61). For healthy children, parents who lived in more disadvantaged areas were less likely to consider their healthy children as healthy (most disadvantaged areas: OR=0.69, 95% CI=0.63-0.76; moderate disadvantaged areas: OR=0.56, 95% CI=0.50-0.62).

Parents who had reported annual household income between $52,000-$103,999 were more likely to perceive their overweight children as healthy ($52,000-$77,999: OR=1.39, 95% CI=1.25-1.54; $78,000-$103,999: OR=1.35, 95% CI=1.22-1.52). Parents who reported an annual household income below $103,999 were twice as likely to perceive their obese children as having a healthy weight ($0-$51,999: OR=2.04, 95% CI=1.82-2.22.; $52,000-77,999: OR=2.38, 95% CI=2.13-2.63).

## Discussion

This study explored the effect of children’s BMI, on the association between parent’s characteristics, SES (assessed by area-level disadvantage and household-level income) and parental perception of their children’s weight status. In this study, approximately 35% of parents misperceived their child’s weight status (23% for healthy weight children; 65% for overweight children and 55% for obese children), a finding consistent with previous studies that a large proportion of parents misperceive their child’s weight status ^5, 9, 15, 19, 33^, with an inclination for underestimation.

Our study results from the adjusted models show that fathers as the primary care providers, parents who completed ≤12 years of school education, and Australian-born parents were more likely to misperceive their child’s weight status. Further, while SES was significantly associated with parental perception of a child’s weight status, the results were inconsistent when measured using area-level disadvantage and household-level income. We found contrasting results for these two SES measures.

### Parents sociodemographic characteristics and perception of their child’s weight status

Most studies on parents’ determinants of misperceptions have mostly included mothers only; hence mothers’ misperception of their child’s weight status is well known with a propensity to underestimate their child’s weight ^5^. However, in line with some other studies ^34, 35^, we found that fathers are more likely than mothers to misperceive their child’s weight status (i.e., fathers are less likely to consider their overweight and obese children as overweight or obese). We do not know nor does the existing literature explain this. Hence, further studies are recommended to understand why fathers compared to mothers are more likely to misperceive their child’s weight status.

Our finding that parents with lower levels of education are more likely to underestimate the weight status of their overweight children is consistent with previous reports^5, 34^ . A recent systematic review, however, found that this evidence was mixed with one-third of studies finding an association and over half finding no association between parental education levels and parental misperceptions of their child’s weight status ^5^.

Further studies are required to understand why Australian-born parents incorrectly perceive their overweight or obese children as healthy weight compared to overseas-born parents. A recent systematic review showed that among the 19 included studies, over half found no association and one-third found associations between ethnicity and parental misperception of their children’s weight status ^5^. As much of the evidence was from the United States of America (USA)^5^, more studies on the effects of ethnicity and parental perceptions of a child’s weight status need to be conducted in other countries.

Literature reviews about the effect of a child’s age, gender and parents’ perception of a child’s weight status suggest that parents generally overestimate their daughter’s weight and underestimate their son’s weight and that parents are less likely to misperceive their child’s weight as children get older ^5^. However, our study results show that the children’s gender and age were not significant determinants of parent’s perceptions of children’s weight status.

### Parents SES and perception of their child’s weight status

Evidence on the association between SES and parental underestimation of their child’s overweight or obese weight status is mixed. Among the 21 studies included in a meta-analysis ^5^, half of the studies found no association; one-third of studies found that parents of low SES are more likely to underestimate their child’s overweight status, and a small number of studies found parents of higher SES are more likely to underestimate their child’s overweight status. It is not clear, however, whether parental misperception was impacted by parents’ SES status, particularly when assessed by household income and area-level disadvantage. We found that parents who lived in the most and middle disadvantaged areas were less likely to perceive their healthy-weight children as having healthy weight, and overweight children as having overweight but more likely to perceive their obese children as slightly overweight or very overweight than parents who lived in less disadvantaged areas. However, we found opposite results when household income was considered as an SES measure. We found that parents who reported the lowest household income (i.e., $0- 51,999) were less likely to consider their obese children as slightly overweight or very overweight but more likely to consider their healthy weight children as healthy weight and overweight children as overweight. Comparing individual and area measures of SES may produce different outcomes although some studies of childhood obesity in Australia have reported similar findings for individual and area SES measures^36^.

### Why do parents misperceive their child’s weight status?

Although overweight and obesity is a global concern, a child’s weight status (healthy or unhealthy) often goes undetected due to many factors. These include a lack of awareness among parents about how to correctly determine their child’s weight status or the need to do so ^37^, increased societal acceptance of children being overweight as it becomes the norm ^19^ and overweight children are typically not perceived to be of concern to parents compared to children who are identified as obese ^19^ especially if the overweight children are physically active and healthy ^38^. Consistent with these studies, our study also found that parents are less likely to consider their child overweight or obese when they do not perceive childhood overweight or obesity as a problem, do not perceive the consumption of sugar drinks as an obesogenic factor, or perceive their child’s overall diet as healthy.

### Effects of parent’s misperception of their child’s weight status

The adverse effects of parental misperception of a child’s weight status are well documented^19, 39, 40^. If parents do not acknowledge their child is at risk for overweight or obesity and its associated complications, it is also less likely that they would take measures to support their children’s healthy lifestyle practices ^10^. Further, misclassification of a healthy child as overweight, obese, or underweight, can lead to negative psychological impacts due to stigmatisation ^19, 40^, reduced interest in physical activity ^41^ and eating disorders ^39^. Hence, routine screening of weight and height is even more necessary and recommended so that parents are aware of their child’s correct weight status and developmental concerns over time.

### Limitations and strengths

There are several limitations of this study. First, this study used the data from parents who self-reported the height and weight of children which may lead to bias ^19^. Parents who did not know their child’s height and weight were called back to give them time to measure their child. Nevertheless, the advantage of our self-reported data is that data collection came from a large representative population sample. In contrast, research with actual measurements from children usually comes from smaller and less representative samples ^15^. Future datasets that include additional characteristics of participants such as parents’ height and weight, children’s perception of their weight status, parents’ BMI, the presence of other overweight or obese children in the family, and characteristics of food and physical environment are warranted. These factors have previously been found to impact parents’ perceptions of their child’s weight status ^5, 19, 37, 42, 43^ and were not measured in this study.

The strengths of this study include a large random sample. Another strength of the study was the information on parental health behaviour perceptions and their association with parent misperception of children’s weight status, such as parents’ perception of overweight and obesity, children’s overall diet, and perceptions of children’s eating behaviour. The inclusion of area-level disadvantage is another strength of this study as this may facilitate population-level interventions. Nevertheless, as explained elsewhere, further studies are needed to understand the discrepancy between different SES measures (household income vs area-level disadvantage) and parent perceptions of their child’s weight status.

## Conclusion

The findings suggest that there may be benefit in designing interventions that focus on improving parents’ misperception of children’s weight status. The groups of parents who should be prioritised for preventive public health interventions to modify the parental misperceptions of their child’s weight status include a) fathers, b) parents who completed ≤12 years of school education and parents who had Trade, TAFE or Diploma educational qualifications, c) parents living in moderately disadvantaged areas and d) parents with low and high household-income. Our findings support the existing strategy that public health interventions need to continue to focus on the education of families and children about healthy weight management, and integration of routine weight and height measurements at population and clinical level settings to help parents develop an accurate awareness of their child’s weight status and overall growth and prevent the risk of chronic disease in the long run. Collecting SES information at both the population and clinical levels would be useful to guide health promotion programs that target healthy weight among children.

## Data Availability

Data available through manuscript

## Acknowledgements

We firstly acknowledge the parents and carers of children living in the South West Sydney Local Health District who generously gave their time to complete the GHK-SWS Survey. We acknowledge the contribution of McNair Yellow Squares who were contracted by the University of Sydney to undertake the survey. We also acknowledge the Growing Healthy Kids Evaluation Advisory Group which contributed to the development of the survey and analysis of the data. Dr Phil Hughes of Engine Group provided the data weighting. We thank Francis Fox for conducting grammar, syntax, and associated edits.

## Authors’ contributions

VK wrote the manuscript and was responsible for the design, drafting, interpretation of data, and extensive revisions of the manuscript. SC contributed to the design, and methodology, conducted analysis of data, wrote the results section, reviewing and revisions of the manuscript. BJ contributed to the design, methodology, results, extensive revisions of the manuscript and oversaw the data analysis and interpretation. JT, MC, MT, and MW contributed to the revisions of the manuscript. All authors read and approved the final manuscript.

